# Creating an e-cohort of individuals with lived experience of homelessness and subsequent mortality in Wales, UK

**DOI:** 10.1101/2020.12.15.20248252

**Authors:** Jiao Song, Charlotte N.B. Grey, Alisha R. Davies

**Affiliations:** Research & Evaluation Division, Knowledge Directorate, Public Health Wales, Cardiff, UK

**Keywords:** homelessness, death, mortality, COVID-19, data linkage, amenable mortality, homeless healthcare, homeless health, precarious housing, insecure housing

## Abstract

**Background:** Homelessness is an extreme form of social exclusion, with homeless people experiencing considerable social and health inequities. Estimates of morbidity and mortality amongst homeless populations is limited due to the lack of recording of housing status across health datasets. The aim of this study is to: (1) identify a homelessness e-cohort by linking routine health data, and (2) explore whether a period of reported past homelessness, places this population at greater risk of morbidity and mortality.

**Methods:** Homelessness identified through linkage across primary, secondary care, and substance misuse datasets in the Secure Anonymised Information Linkage (SAIL) Databank. Mortality was examined through linkage to the Office for National Statistics mortality data.

**Results:** E-cohort of 15,472 individuals with lived experience of homelessness identified. Of those, 21 individuals died between February and July 2020 from COVID-19. Those with lived experience of homelessness had increased mortality from many causes including COVID-19, i.e. accidents, liver diseases and suicides.

**Conclusion:** Linking multiple routine datasets provides a more comprehensive dataset of a marginalised population. Application of the cohort demonstrated that individuals with lived experience of homelessness have increased mortality from COVID-19 and other causes. The underlying reasons, health needs, and causes of death warrant further exploration.

**Thumbnail Sketch**
**What is already known on this subject?**
- Homelessness includes the street homeless and other precarious/insecure housing situations (e.g. sofa surfing), and this population suffer multiple health and social inequalities, known to have a detrimental impact on short and longer-term health outcomes
- Understanding the health impact resulting from lived experiences of homelessness is limited due to a lack of consistent and accurate definition and recording of homelessness in health and social care datasets
- Homelessness policy in Wales during the COVID-19 pandemic supported the current street homeless, but what less is understood on the impact of those with a period of reported homelessness and insecure housing situations prior to COVID-19

**What this study adds?**
- This study has created an e-cohort of 15,472 individuals with lived experience of homelessness recorded by NHS Wales services between since 2014 in Wales, from routinely collected primary care, secondary care, and substance misuse datasets stored in the Secure Anonymised Information Linkage (SAIL) Databank
- This demonstrates it is possible to link multiple routinely collected health and care datasets to create a reproducible e-cohort that can rapidly be used (prospectively and retrospectively) to support research and policy
- Our methodology allows for a more sensitive definition of those in current and past vulnerable housing circumstances that will allow for improved understanding of health impacts in this group, and inform practice and policy response
- Using the e-cohort we examined the relationship between homelessness and mortality outcomes during the COVID-19 pandemic and found individuals with lived experience of homelessness, even if resolved, were at greater ongoing risk of mortality. The underlying reasons, health needs, and causes of death in this population warrants further exploration.

## Introduction

Social inequalities have been observed in historical pandemics (1), and emerging evidence for the COVID-19 (SARS-CoV-2) pandemic suggests a similar disproportionate impact on the already disadvantaged (1,2). Contributing factors include underlying inequalities in the social determinants of health and higher prevalence of underlying chronic disease (1). Additionally, the short and long term political and economic consequences of the virus are likely to disproportionately impact the health of the already vulnerable (1), including contributing to housing insecurity resulting in new threats of homelessness (3).

Homelessness is a complex and persistent public health challenge (4,5) and extreme form of social exclusion (6,7) affecting at least 7% of the Welsh adult population (8), with this population having a higher proportion of multiple, complex health needs (9–11). Homelessness can be defined as having either a lack of adequate housing or living in housing below a minimum adequacy standard (12), and can be chronic, episodic, or transitional (6,13). Homelessness therefore includes both the street homeless but also those with precarious or insecure living arrangements (5,9,13,14).

The homeless population experience extreme health inequities (7), with ill-health being both a cause and consequence of homelessness (15). A high proportion of multiple health issues including poor mental health, physical illness, or substance misuse is seen in this group (9–11). Data from England suggests that homeless individuals are four times more likely to use Accident and Emergency services (A&E) than the general population, and this over-representation in unscheduled care costs the NHS eight times more than the general population (16–18). Excess mortality in this population is significantly higher than in the general population (7,17,19), with data from England and Wales showing that the mean age of mortality is over 30 years younger in men and women compared to the general population (19). During COVID-19, initial estimates up until the end of June 2020 suggested that for England there were 16 deaths in the currently homeless population where COVID-19 was mentioned on the death certificate, either as an underlying cause or a contributory factor, and none in Wales (20). Yet a lack of reliable data on housing status in routine health and care data is likely to underestimate both the extent of precarious housing status and homelessness, and the resulting short and long term impact on health (5,21).

The aims of this study is to generate a population-based retrospective e-cohort of people with lived experience of homelessness, using linked routinely collected health data; and to apply that cohort to examine the relationship between homelessness and mortality during the COVID-19 pandemic.

## Methods

We adopted a broad definition of lived experience of *homelessness* to include the street homeless and those with precarious or insecure housing circumstances (12).

### Cohort construction

All patients in Wales who accessed NHS health services for any routine activity at any point since 2014 *and* had a homelessness/precarious housing code in one or more of four datasets held in the Secure Anonymised Information Linkage (SAIL) Databank were flagged as having *lived experience of homelessness* and included in our e-cohort (Figure 1). The SAIL Databank assigns an anonymous linkage field unique for each individual using a privacy-protected split file approach, which can then be used to link across datasets at individual level (22,23). The clinical codes and patient status identifying *homelessness* in routine data (Figure 1) was drawn from 11 expert clinicians, academic researchers, and stakeholders (listed in the Acknowledgements).

**Figure 1:**
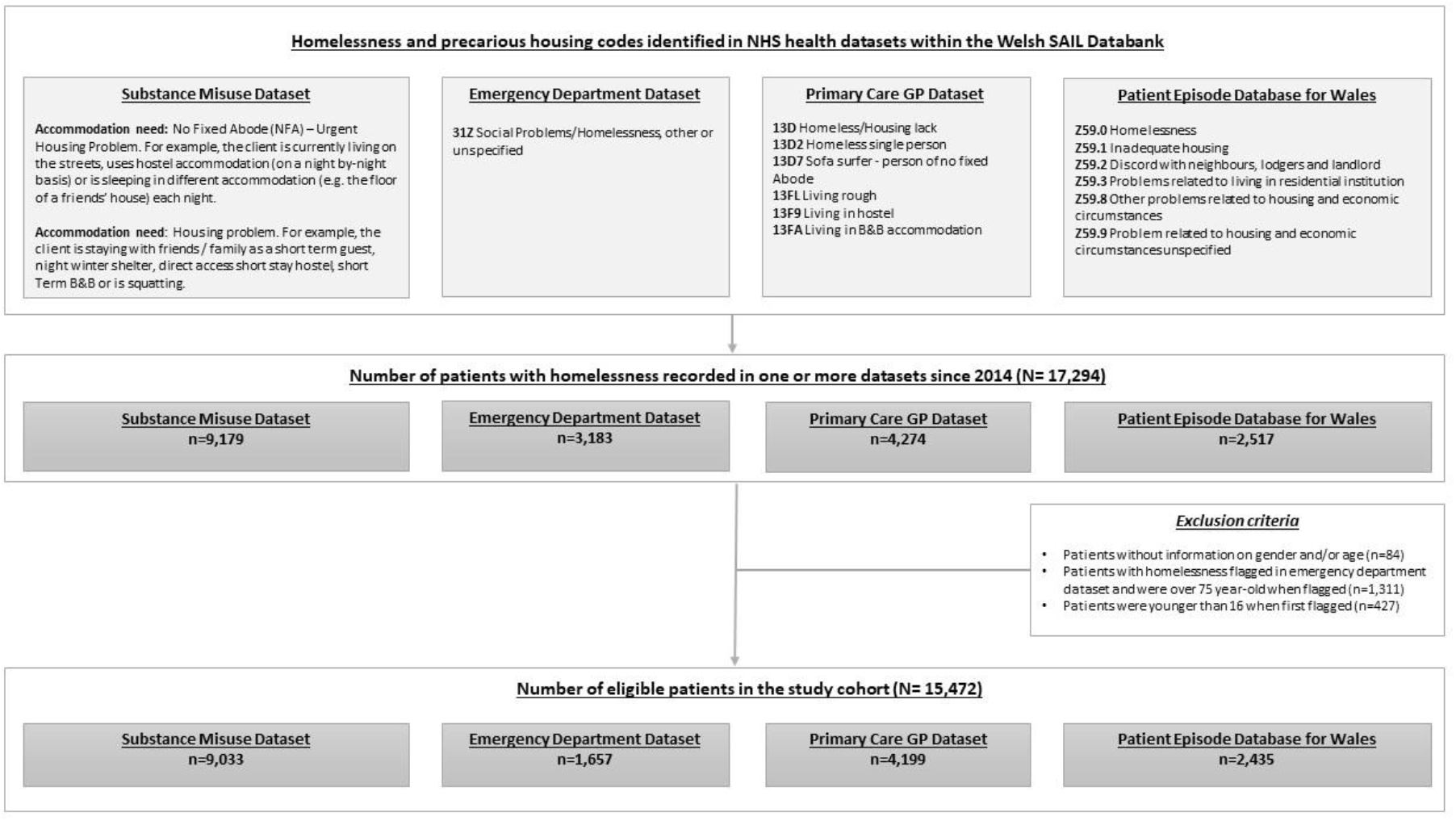
Flow diagram of the construction of our e-cohort of individuals with lived experience of homelessness.

The study period started in 2014, when the Housing (Wales) Act 2014 came into force. Age was assigned when individuals appeared in the cohort for the first time.

### Exclusions

Patients without demographic information (age and/or gender) were excluded. Patients aged >74 years and *<u>only</u>* flagged in the Emergency Department dataset with diagnosis code 31Z (definition: social Problems/Homelessness, other or unspecified) were also excluded. This was necessary as it was not possible to distinguish homelessness and social care needs that might be associated with older age (24). Patients <15 years were also excluded.

### Analysis

We used SQL Db2 to interrogate data in the SAIL Databank and performed the analysis using R for Windows (www.r-project.org).

### Ethics statement

This study is based on anonymised routinely collected electronic health records. All routinely collected anonymised data held in SAIL Databank are exempt from consent due to the anonymised nature of the databank (under section 251, National Research Ethics Committee (NREC)). We have applied and been granted approval by the independent Information Governance Review Panel (IGRP) for permission to conduct this study (project number 0968). The IGRP contains independent members from NREC and British Medical Association (BMA), as well as lay members. The review process has checked that the study we are is useful, not service evaluation, and will not break anonymisation standards.

## Results

### Cohort characteristics

We identified 15,472 individuals with lived experience of homelessness in Wales at any time point since 2014. Of these, 10,609 (68.6%) were male. At the first time they appeared in the cohort, 7,107 (45.9%) were between 16 and 34 years old and 462 (3.0%) were above 75 (Table A1, Supplementary Material).

### Deaths between January 2014 and May 2020

We linked our e-cohort with mortality data (until July 2020) from the Office for National Statistics (ONS Deaths) via the SAIL Databank. 1,286/15,472 (8.3%) of our e-cohort died between January 2014 and July 2020. Of these, 882/1,286 (68.6%) were male and 335/1,286 (26.0%) were under 45 years old when they died.

We compared the underlying causes of deaths (25) for deaths registered for the three cohorts listed below (Figure 2 and Table A2, Supplementary Material).

**Figure 2:**
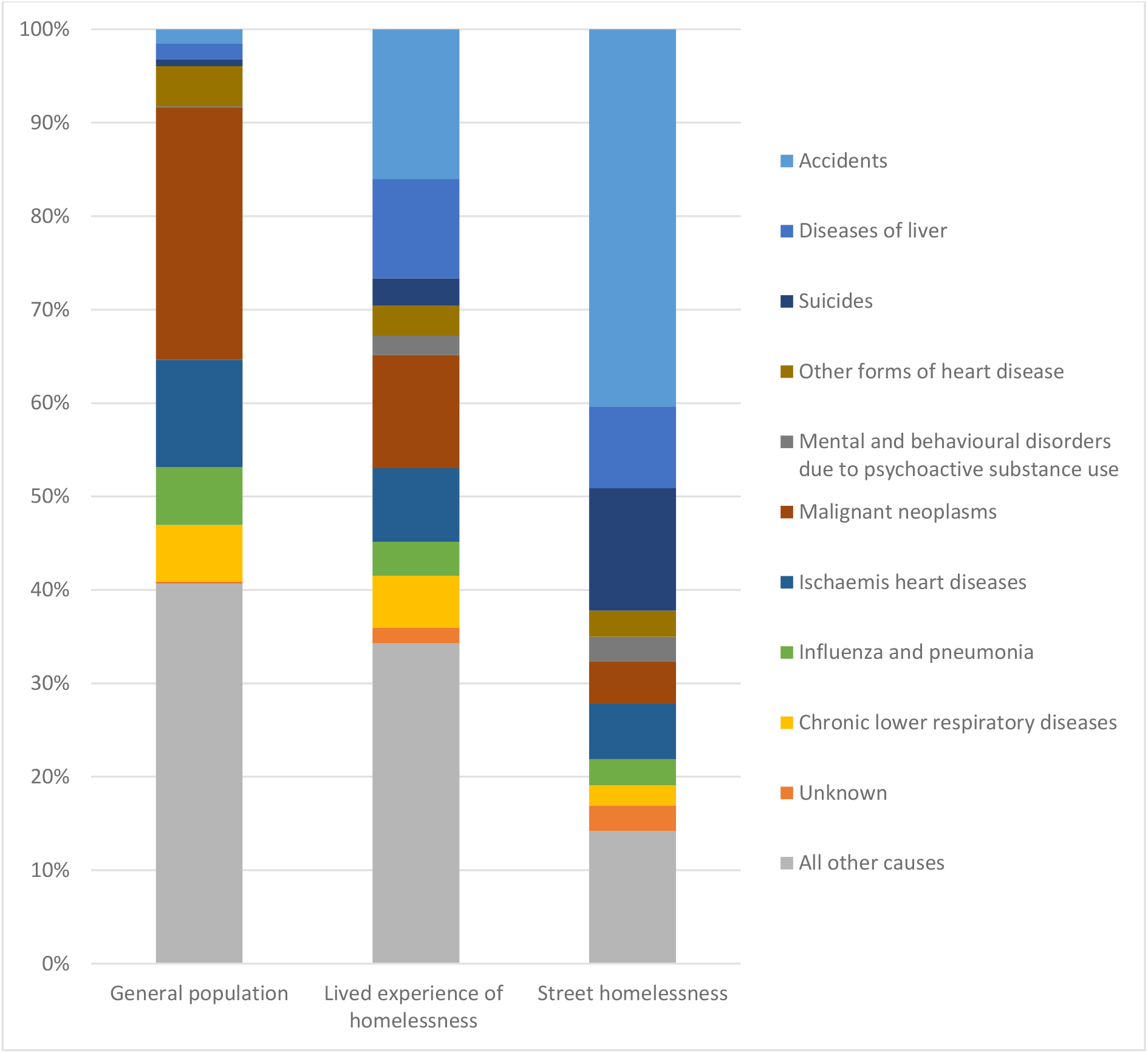
Comparison on underlying causes of death between three cohorts.

- General population mortality data for Wales since 2014(ONS Deaths)
- Our e-cohort: *lived experience of homelessness* since 2014 in Wales
- Street homelessness in England and Wales (using estimated deaths of homeless people by underlying causes of death in England and Wales 2017 as a proxy (19))

Figure 2 demonstrates that our e-cohort of those with lived experience of homelessness differs to both the general population and the street homeless cohorts. In our e-cohort, deaths from accidents and suicides, and mental and behavioural disorders due to psychoactive substance use are higher than the general population, but less than in the street homeless. The proportion of deaths due to liver disease were greatest amongst our e-cohort, compared to the general population and the street homeless.

### Deaths during the COVID-19 Pandemic in those with lived experience of homelessness

The first confirmed case of coronavirus in the UK was in January 29, 2020. Between February and July 2020, 141 deaths occurred within our e-cohort. 101/141 (71.6%) of the deaths were in males. 16/141 (11.3%) were under 45 years old when they died. 21/141 (14.9%) deaths were recorded with COVID-19 as the underlying (primary) cause of death comparing to 13.9% in general population.

## Discussion

Our study created a unique e-cohort of individuals across Wales with recorded lived experience of homelessness from multiple NHS health datasets that can be quickly used to analyse health needs and mortality in this population, as demonstrated here for the COVID-19 pandemic. A key strength is the production of an e-cohort bringing together data from multiple databases, compared to previous studies (26), providing a more comprehensive assessment of the numbers having lived experience of homelessness in Wales. The method used to identify the e-cohort is reproducible and can be applied to health care data over time to generate a comprehensive understanding of the health care needs of those with lived experience of homelessness and inform service planning and policy. This cohort can also help to evaluate future initiatives to support this vulnerable population in Wales.

Compared to similar data linkage studies (26), we use both a broader definition of homelessness that considers precarious and insecure housing status, alongside street homelessness, and did not restrict homelessness as recorded by single care provider at one time point. Whilst this is less specific than street homelessness alone, it does capture those with precarious housing experiences and the longer-term impact of lived experience of homelessness. Our results suggest that this more diverse group of those with *lived experience of homelessness* have poorer health outcomes than the general population, but not as poor as those street homeless. This is an important consideration for health and care, where early identification of those in precarious housing situations may help better inform integrated support addressing health needs and social circumstances to reduce the health inequalities.

Previous attempts to understand mortality in the street homeless population during COVID-19 was undertaken that found zero cases of COVID-related deaths in Wales (19). However, from our data, we identified 141 individuals with *lived experience of homelessness* in Wales who died during COVID-19, 21 of whom died with COVID-19 as the primary cause of death. This suggests that those who have experienced homelessness are likely to have poorer health than the general population, combined with complex longer term mental and physical health needs (8) amplifying the prevalence and severity of COVID-19 (1,27).

Nonetheless, there are a number of limitations to the approach taken. A potential source of systematic bias is that how often/well the clinical codes were used in the datasets is unknown. We will explore that further in future analyses. We do not know the extent of the potential barriers this group have accessing healthcare services and whether there is a potential underestimation of the problem. We plan to further explore the differences in causes of death in this group, considering the impact of comorbidities on mortality. Another source of bias is that currently primary care data coverage in SAIL Databank sits at around 80% in Wales.

## Data Availability

This study is based on anonymised routinely collected electronic health records held in Secure Anonymised Information Linkage (SAIL) Databank (Project ID 0968).

## We would like to thank the following expert clinicians and stakeholders for their contribution in scoping and reviewing how to capture homelessness in routine data

Dr Simon Braybrookand and Dr Kay Saunders (Butetown Medical Practice); Hannah Browne-Gott, Dr Peter Mackie, Dr Naomi Stanton and Dr Ian Thomas (Cardiff University); Asim Butt (Mortality, Office for National Statistics); Rebecca Jackson (Shelter Cymru); Adam Golten and Alex Osmond (The Wallich); Judith David (Environment, Sustainability and Housing Stats Welsh Government).

### This study makes use of anonymised data held in the Secure Anonymised Information Linkage (SAIL) Databank

We would like to acknowledge all the data providers who make anonymised data available for research.

## Supplementary Material

**Table A1:** E-cohort demographics.

| Age group | Gender |  |
| --- | --- | --- |
|  | Male | Female |
| 16-34 | 4741 | 2366 |
| 35-44 | 2712 | 1027 |
| 45-54 | 1770 | 639 |
| 55-64 | 752 | 328 |
| 65-74 | 416 | 259 |
| 75+ | 218 | 244 |

**Table A2:** Underlying causes of death between three cohorts.

| <u>Underlying causes of death</u> | Lived experience of homelessness |  | General population |  | Street homelessness |  |
| --- | --- | --- | --- | --- | --- | --- |
|  | Counts | % | Counts | % | Counts | % |
| Accidents | 206 | 16.0% | 3345 | 1.5% | 241 | 40.4% |
| Chronic lower respiratory diseases | 72 | 5.6% | 13265 | 6.0% | 13 | 2.2% |
| Diseases of liver | 137 | 10.7% | 3646 | 1.7% | 52 | 8.7% |
| Influenza and pneumonia | 47 | 3.7% | 13668 | 6.2% | 17 | 2.8% |
| Ischaemic heart diseases | 101 | 7.9% | 25135 | 11.5% | 35 | 5.9% |
| Malignant neoplasms | 156 | 12.1% | 59321 | 27.0% | 27 | 4.5% |
| Mental and behavioural disorders due to psychoactive substance use | 26 | 2.0% | 258 | 0.1% | 16 | 2.7% |
| Other forms of heart disease | 42 | 3.3% | 9415 | 4.3% | 17 | 2.8% |
| Suicides | 37 | 2.9% | 1640 | 0.7% | 78 | 13.1% |
| Unknown | 21 | 1.6% | 501 | 0.2% | 16 | 2.7% |
| All other courses | 441 | 34.3% | 89263 | 40.7% | 85 | 14.2% |
| Total | 1286 | 100.0% | 219457 | 100.0% | 597 | 100.0% |

